# PySpiro: A Python Library for Lung Function Reference Equations and Severity Classification

**DOI:** 10.64898/2026.10.06.26364407

**Authors:** Hendrik Pott, Bernd Schmeck, Dominik Heider, Roman Martin

**Author notes:** **Corresponding author details:** Name: Dr. Roman Martin, Address: Institute of Medical Informatics, University of Münster, Albert-Schweitzer-Campus 1, 48149 Münster, Germany.

## Abstract

**Background and Objective:** Lung function reference equations are essential for interpreting spirometry and related modalities, yet existing tools are fragmented, modality- and language-specific (predominantly R), and lack integrated severity classifiers. We present PySpiro, an open-source Python library unifying 23 spirometry, 1 multiple breath washout, 2 diffusing capacity, 1 static lung volume, and 2 oscillometry reference equations under a single API, with 10 clinical severity classifiers.

**Methods:** PySpiro was implemented in Python 3 using NumPy and pandas, with all equations coded directly from literature. The unified API exposes predicted medians, z-scores, limits of normal (LLN/ULN), and raw Lambda-Mu-Sigma (LMS) triplets per parameter, plus a vectorised compute() method for batch processing at the DataFrame level. Unit-tests covering every equation and classifier verify agreement with published values and confirm correct boundary, out-of-range, and missing-input handling.

**Results:** PySpiro covers spirometry equations from 1971 to 2022 across Caucasian, Asian, Indian, and race-neutral global populations, plus non-spirometry modalities (multiple breath washout, diffusing capacity, static lung volumes, oscillometry). Classifiers span COPD, interstitial lung disease, pattern and severity grading, bronchodilator reversibility, and primary ciliary dyskinesia. Compared with rspiro (5 equations, 4 spirometry) and the only prior Python package (spiref, 2 equations, no z-scores or % predicted), PySpiro offers substantially broader coverage, multi-modality support, a richer output API, and 10 classifiers absent from both.

**Conclusions:** PySpiro is freely available (PyPI: pyspiro; doi:10.5281/zenodo.15519193) and provides a comprehensive, validated, and extensible platform for lung function data analysis in research pipelines.

## 1. Introduction

Spirometry is the most important test of pulmonary function. It is paramount to the diagnosis and monitoring of both obstructive and restrictive lung diseases, e.g. chronic obstructive pulmonary disease, the third leading global cause of death. Interpreting spirometric measurements requires comparison against population-specific reference values, which are calculated by reference equations and typically expressed as a percentage of the predicted median or as a z-score relative to the reference distribution [1,2]. Over the past five decades, a landscape of more than 30 of these reference equations has been published, each developed in different populations and age ranges, and using different statistical methodologies [1]. Moreover, modern lung function laboratories may complement spirometry with measurement of diffusing capacity for carbon monoxide (DLCO), static lung volumes, multiple breath washout (MBW) with calculation of the lung clearance index (LCI), and oscillometry (impulse oscillometry systems, IOS). However, each of these modalities has its own population-specific reference equations, and the lack of unified software tools forces researchers and laboratories to work with fragmented, equation-specific implementations.

The most widely used tool for computing reference values is rspiro [3], an R package with at least 36,429 downloads to date [4]. It implements four spirometry equations (GLI-2012, the race-neutral GLI-2022, NHANES III, and JRS-2014) together with the GLI-2017 diffusing-capacity equation. Moreover, rspiro covers only a small subset of published spirometry equations, is implemented in R only, provides no upper limits of normal, and contains no clinical severity classifiers. The only comparable Python package, spiref [5], implements just two equations and returns only predicted values and lower limits of normal, without z-scores, percentage of predicted, upper limits of normal, or DataFrame support. Neither package covers multiple breath washout, static lung volumes, or oscillometry, and neither provides severity classifiers.

Python has become the dominant programming language in scientific computing, data science, and clinical informatics pipelines. The absence of a comprehensive, validated Python library for lung function reference equations represents a significant gap, detrimental to translational respiratory research. To address this, we developed PySpiro, an open-source Python package that:

1. Implements 23 spirometry reference equations spanning 1971–2022, including the three most widely recommended current equations [1,6,7] and equations for populations underserved by existing tools (Indian, Korean, Japanese, Chinese populations);
2. Extends beyond spirometry to cover MBW [8], DLCO [9,10], static lung volumes [11], and oscillometry [12,13];
3. Exposes a unified, consistent API: percent(), zscore(), lln(), uln(), lms(), all(), and compute(), across all equations and modalities;
4. Provides 10 clinical severity classifiers for COPD, ILD, lung function pattern and severity, bronchodilator reversibility, primary ciliary dyskinesia, and COPD exacerbations;
5. Integrates with pandas DataFrames for batch processing of research datasets.

## 2. Methods

### 2.1 Implementation

PySpiro is written in Python 3 and builds on NumPy and pandas, with matplotlib and scipy as optional dependencies for visualisation. Each equation was implemented directly from its primary publication if accessible, otherwise from secondary literature, with coefficients taken from the publications or their supplementary materials. A small hierarchy of base classes factors out the shared mathematics so that each equation only needs to supply its own predicted values, while the same scalar API is exposed across every equation and modality (**Figure 1**). Inputs that fall outside an equation’s validated age or height range are handled by a configurable strategy and, by default, return a missing value rather than an extrapolated one. Full details of the class architecture, coefficient storage, interpolation, limit-of-normal derivation, and out-of-range handling are given in the Supplementary Methods.

**Figure 1.**
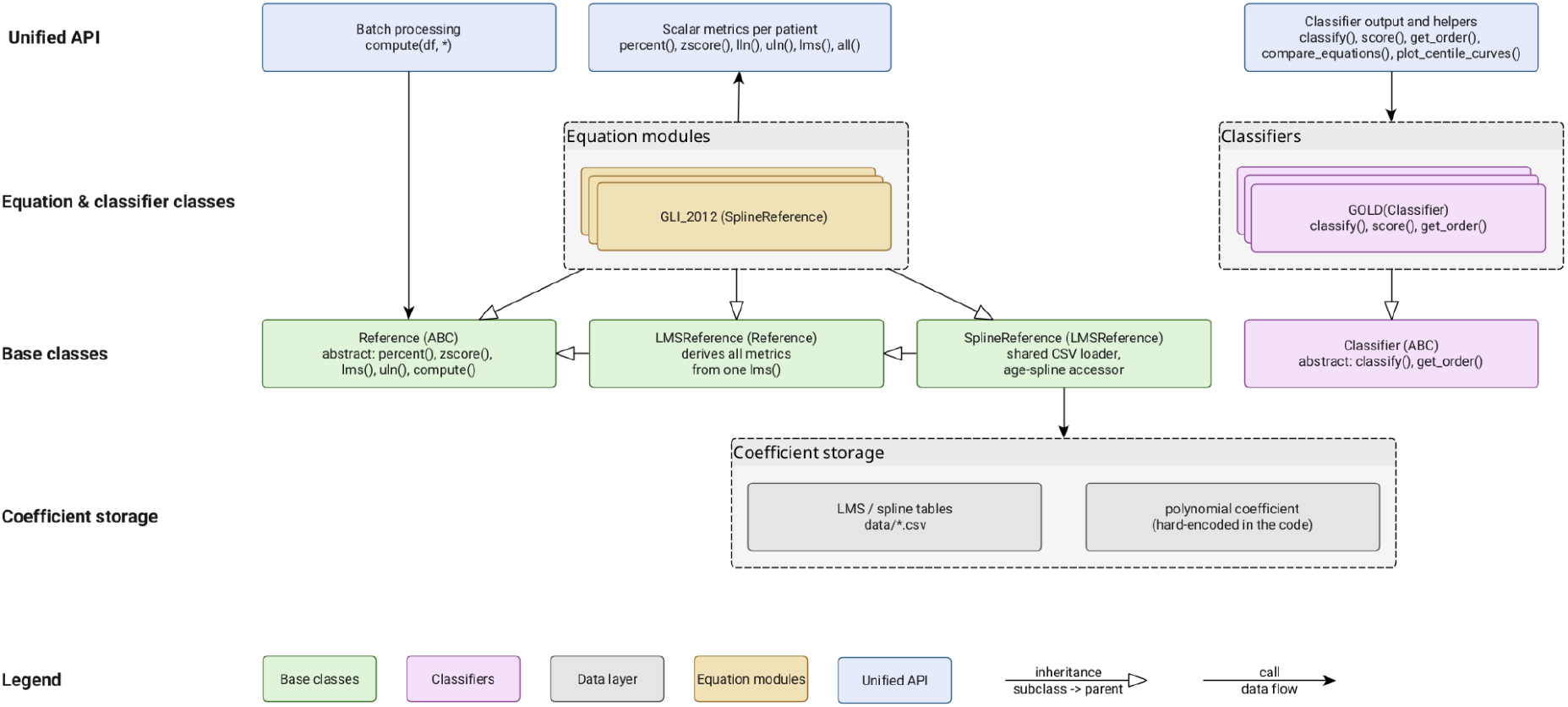
PySpiro software architecture, organised in four layers: coefficient and spline tables, shared base classes, the equation and classifier modules, and the unified API on top.

### 2.2 Equations and severity classifier coverage

A comprehensive list of all 23 implemented spirometry equations and their primary publications is listed in **Table 1**. The most commonly used current equations are GLI_2012 [1] and BOWERMAN_2022 [7]. Both use the GAMLSS LMS methodology and are fully supported including z-scores, LLN, and ULN. HANKINSON_1999 [6], the standard for the United States, is implemented using the lower-limit-of-normal equations published in the original paper. Where polynomial-era equations instead provide a residual spread term, the lower limit of normal is derived as the predicted value minus 1.645 times that term. For completeness, **Table 2** lists all reference equations beyond pure spirometry.

**Table 1.** Spirometry reference equations implemented in PySpiro, in reverse chronological order. For each equation the reference population, validated age range, and availability of a lower limit of normal and z-score are shown; where the age range differs by sex, the wider range is given. A tick in the LLN / z-score column indicates that the equation provides a published or derivable limit of normal, enabling both z-score and LLN output; a dagger (†) indicates that an LLN is available but no z-score; equations marked with a dash return predicted values and percent predicted only.

| PySpiro class | Population | Age range | LLN / z-score | Ref. |
| --- | --- | --- | --- | --- |
| BOWERMAN_2022 | Race-neutral (GLI global) | 3–95 y | ✓ | [7] |
| AGARWAL_2020 | Western Indian (rural Pune) | 20–80 y | ✓ | [14] |
| JO_2018 | Korean (KNHANES IV–V) | 19–90 y | ✓ | [15] |
| JIAN_2017 | Han Chinese | 4–80 y | ✓ | [16] |
| DESAI_2016 | Western Indian (Mumbai) | 18–82 y | ✓ | [17] |
| KUBOTA_2014 | JRS (Japanese) | 17–95 y | ✓ | [18] |
| CHHABRA_2014 | Northern Indian | 18–71 y | ✓ | [19] |
| GLI_2012 | Multi-ethnic | 3–95 y | ✓ | [1] |
| KUSTER_2008 | Swiss LuftiBus | 18–80 y | † | [20] |
| CHOI_2005 | Korean (KNHANES) | 18–80 y | - | [21] |
| HANKINSON_1999 | NHANES III (US) | 8–80 y | ✓ | [6] |
| QUANJER_1995 | White European children | 6–18 y | - | [22] |
| WANG_1993 | White & African-American children (US) | 6–18 y | - | [23] |
| ECCS_1993 | European Caucasian | 18–70 y | - | [24] |
| ROBERTS_1991 | White urban (UK) | 18–86 y | - | [25] |
| ZAPLETAL_1987 | European children | 6–18 y | - | [26] |
| KNUDSON_1983 | Caucasian (US) | 6–90 y | - | [27] |
| CRAPO_1981 | Caucasian (US), non-smokers | 15–91 y | - | [28] |
| WARWICK_1980 | Caucasian children (US) | 0–18 y | - | [29] |
| HSU_1979 | White, African-American, Mexican-American | 7–20 y | - | [30] |
| MORRIS_1971_1973 | Caucasian (US), non-smokers | 20–90 y | - | [31,32] |
| CHERNIACK_1972 | Caucasian (US) | 15–79 y | - | [33] |
| POLGAR_1971 | Children | 4–17 y | - | [34] |

**Table 2.** Reference equations beyond spirometry implemented in PySpiro. † SCAPIS_2023 is the only implemented equation providing post-bronchodilator reference values, spanning both spirometry and diffusing capacity. * CALOGERO_2013 is height-based (≈92–159 cm) rather than age-based.

| Functionality | PySpiro class | Population | Age range | Variables | Ref. |
| --- | --- | --- | --- | --- | --- |
| MBW | RAMSEY_2024 | GLI global | 2–80 y | LCI, FRC | [8] |
| DLCO | GLI_2017 | Caucasian | 5–80 y | DLCO, KCO | [9] |
| Spirometry + DLCO † | SCAPIS_2023 | Swedish | 50–65 y | pre-/post-BD FEV<sub>1</sub>, FVC, FEV<sub>1</sub>/FVC; post-BD DLCO, KCO | [10] |
| Static volumes | GLI_2021 | European ancestry | 5–80 y | TLC, RV, FRC, VC | [11] |
| Oscillometry | SCHULZ_2013 | KORA Germans | 40–65 y | R10, R15, R25, R35, X10, X20, X25, X35 | [12] |
|  | CALOGERO_2013 | Caucasian children | * | Rrs6, Rrs8, Rrs10, Xrs6, Xrs10, AX, Fres | [13] |

Reference values tell you where a measurement sits relative to a healthy population, but clinical decisions usually depend on a further step: turning those values into a pattern, a stage, or a severity grade. PySpiro therefore ships ten classifiers alongside the equations, each implemented from a published guideline, consensus standard, or peer-reviewed scoring system. Together they cover obstructive and restrictive pattern recognition, lung-function severity grading, bronchodilator reversibility, COPD staging and exacerbation severity, idiopathic pulmonary fibrosis staging, and screening for primary ciliary dyskinesia. **Table 3** lists each classifier with the inputs it expects and the categories it returns.

### 2.3 Validation

Numerical accuracy was verified by a comprehensive unit-test suite, implemented with the Python unittest framework, covering every equation and classifier and including boundary, out-of-range, and missing-input handling. For each equation, tests were written against one or more of the following sources: (a) worked examples from the primary publication; (b) independently computed values using the published coefficient tables; (c) mathematical identities (e.g., that the z-score at the predicted median equals 0; that the z-score at the LLN equals −1.645). All tests are run in a continuous integration environment against every commit.

**Table 3.** Clinical severity classifiers implemented in PySpiro. For each classifier the table lists the inputs it consumes, the categories or scores it returns, and the guideline, consensus standard, or scoring system it implements. Each classifier is a direct encoding of the published decision rule and operates on the outputs of the reference equations (for example, z-scores or percent predicted) together with any additional clinical inputs the rule requires.

| Classifier | Basis / inputs | Output | Ref. |
| --- | --- | --- | --- |
| ATS_ERS_2022 | FEV<sub>1</sub>/FVC, FVC, FEV<sub>1</sub>, TLC z-scores | Normal / Obstructive / Restrictive / Mixed / Non-specific | [2] |
| LF_SEVERITY_2022 | Any lung function z-score (parameter-agnostic) | Normal / Mild / Moderate / Severe | [2] |
| BDR_2022 | Pre/post FEV<sub>1</sub> and/or FVC vs predicted | Positive (FEV<sub>1</sub>) / Positive (FVC) / Positive (both) / Negative | [2] |
| GOLD | FEV<sub>1</sub> % predicted (COPD) | Stage 1–4 | [35] |
| GOLD_ABE | Exacerbation history + CAT / mMRC (COPD) | Group A / B / E | [35] |
| STAR | FEV<sub>1</sub>/FVC ratio (COPD) | Stage 1–4 | [36] |
| BODE | BMI + FEV<sub>1</sub> % predicted + mMRC + 6MWT (COPD) | Score 0–10, quartiles 1–4 | [37] |
| GAP | Sex + age + FVC % predicted + DLCO % predicted (IPF) | Score 0–8, stages 1–3 | [38] |
| WODEHOUSE_2003 | Nasal nitric oxide (nNO, ppb) | PCD range / Normal | [39] |
| ECOPD_ROME_2021 | Dyspnoea VAS, RR, HR, SaO<sub>2</sub>, CRP | Mild / Moderate / Severe | [40] |

As an additional, independent check, every reference equation that PySpiro shares with the established rspiro R package was cross-validated numerically against rspiro, both on a synthetic cohort of 5,000 patients and on the real publicly available lNHANES 2011–2012 cohort (6,309 participants), with agreement assessed by Bland–Altman analysis. Where its narrower output allowed, the Python package spiref was included in the same comparison.

### 2.4 Runtime benchmarks

To characterise computational performance, we benchmarked PySpiro’s vectorised compute() interface against the rspiro R package and the spiref Python package across cohort sizes ranging from 100 to 100,000 patients. Each package was timed on identical patients on a single core, and the median of three runs is reported. For PySpiro and rspiro we measured z-score computation time for five representative equations; because spiref computes only predicted values, it was benchmarked separately on the predicted-value task it shares with the other two packages. As the three packages differ in how they are invoked, the reported throughput reflects each tool’s native calling interface as much as the underlying equation arithmetic.

## 3. Results

### 3.1 Comparison with existing tools

Across the two other openly available reference-equation tools, PySpiro offers substantially wider coverage. It implements 23 spirometry equations against 4 in rspiro and 2 in spiref, and is the only one of the three to extend beyond spirometry, adding six equations spanning multiple breath washout, diffusing capacity, static lung volumes, and oscillometry (rspiro provides one, the GLI-2017 diffusing-capacity equation; spiref none). Its output API is also richer: PySpiro exposes all common metrics including z-scores, percent predicted, and both limits of normal, whereas spiref returns only predicted values and the lower limit of normal, and neither comparator reports an upper limit of normal or offers cross-equation comparison and centile visualisation. The ten clinical classifiers are absent from both. A full feature-by-feature breakdown, covering implemented equations, modalities, output metrics, and software characteristics such as language, licence, and package manager, is given in **Tables S1a** to **S1d**.

### 3.2 Validation against published reference values

To confirm that each equation was implemented correctly, we compared PySpiro’s output against the values reported in the original publications, drawing on worked examples and coefficient tables from the source papers wherever they were available. **Table S2** collects these checks for a representative set of equations and parameters. In every case the computed values matched the published figures to the precision reported in the source.

### 3.3 Cross-validation against existing implementations

Beyond agreement with published values, we confirmed that PySpiro reproduces an independent implementation by comparing it directly with the rspiro R package. On a synthetic cohort of 5000 adults spanning every shared equation, agreement was excellent throughout: the LMS-based equations matched to spline-interpolation precision (z-score bias < 5×10^−3^, 95% limits of agreement within ±0.02 z-units) and JRS-2014 agreed to machine epsilon, with the only larger residuals confined to the NHANES III child/adult coefficient transition, where bias nonetheless remained essentially zero. We then repeated the comparison on real measurements from the NHANES 2011–2012 cohort (6309 participants). Agreement on real data matched the synthetic result: NHANES III values were bit-identical between the two packages for 98.2% of evaluations, the exceptions being female participants aged 18–19 y, for whom the two packages apply the child/adult coefficient cut-off at slightly different ages, while GLI-2012 agreed to spline-interpolation rounding (largest difference 6×10^−4^). On this cohort PySpiro also computed the full set of reference values at a throughput comparable to vectorised R. The Python package spiref, where its more limited output permitted comparison, likewise agreed with both packages. **Figure 2** shows the Bland–Altman agreement on the real NHANES cohort; the synthetic-cohort cross-check across all shared equations and full agreement statistics are given in the Supplementary Material (**Tables S3–S4, Figure S1**).

**Figure 2.**
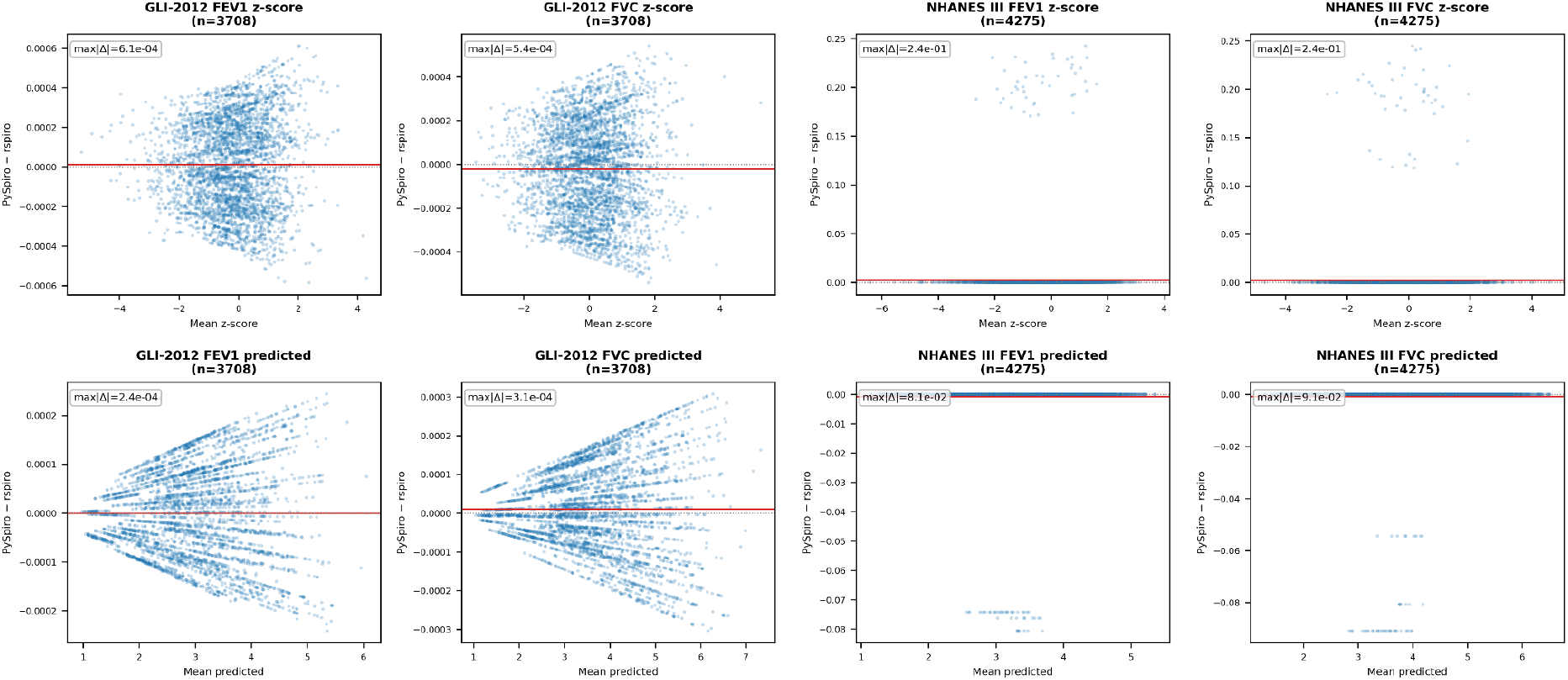
Bland–Altman agreement between PySpiro and rspiro on real NHANES 2011 to 2012 participants (6309 retained; equation-specific subsets shown per panel). Panels cover the two shared equations for each parameter (FEV_1_, FVC) and each metric, with z-scores in the top row and predicted medians in the bottom row; each plots the per-patient difference (PySpiro minus rspiro) on the y-axis against the mean of the two estimates on the x-axis. In every panel the solid line marks the mean difference (bias) and the dotted line zero, with the maximum absolute difference annotated. The GLI-2012 panels show a tight scatter at the 10^−4^ level, reflecting spline-interpolation rounding, while the NHANES III panels are pinned at zero (bit-identical agreement) apart from a sparse set of outliers, all female participants aged 18 to 19 y, where PySpiro and rspiro apply the child/adult coefficient cut-off at slightly different ages.

### 3.4 Runtime benchmark

Across cohort sizes from 100 to 100,000 patients, runtime scaled linearly with cohort size for all three packages, and reference-value computation never became a practical bottleneck. PySpiro processed roughly 10,000 to 15,000 patients per second on a single notebook CPU core, completing a 10,000-patient cohort in about a second. For the LMS-based equations rspiro’s vectorised R routines ran a few times faster than PySpiro’s row-wise evaluation, while the two were comparable for the NHANES III polynomial; for the predicted-value task that spiref also supports, spiref’s lightweight list-comprehension approach achieved the highest throughput, with rspiro’s vectorised routines also exceeding PySpiro’s row-wise evaluation; the reported figures reflect each tool’s native calling convention as much as the underlying arithmetic. Full timings and throughput figures are given in **Figure 3** and **Tables S5** to **S7**.

**Figure 3.**
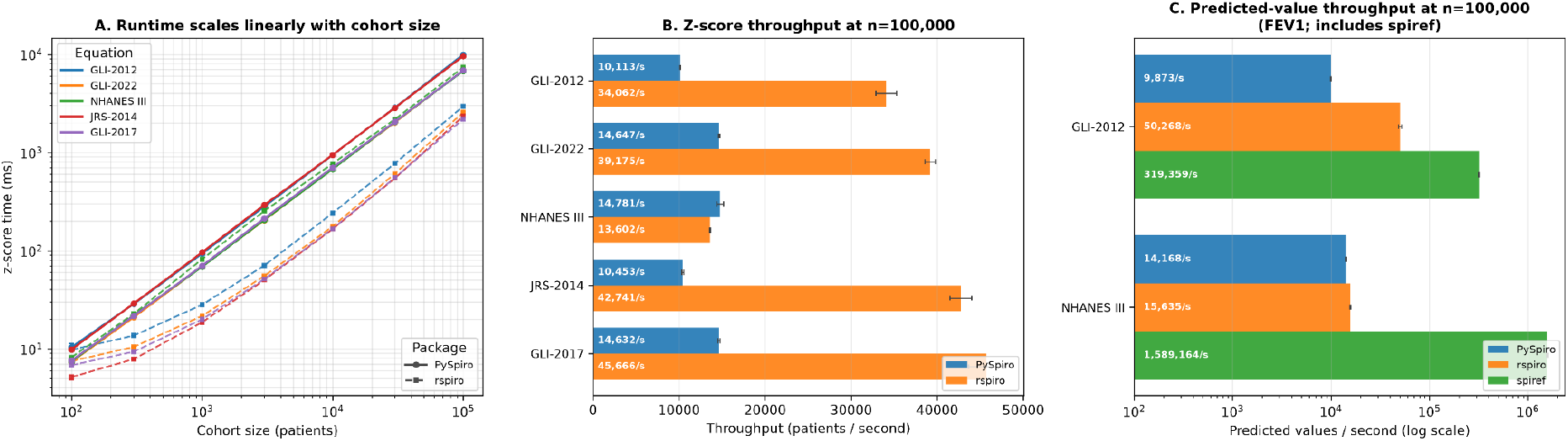
Runtime benchmark of PySpiro against rspiro and spiref. **(A)** z-score computation time across cohort sizes for five representative equations, scaling linearly with cohort size for both PySpiro and rspiro. **(B)** z-score throughput at 100,000 patients: rspiro is faster for the LMS-based equations, while the two are comparable for the NHANES III polynomial. **(C)** predicted-value throughput at 100,000 patients for the single task spiref supports; the large throughput differences reflect calling convention (spiref’s list comprehension vs. PySpiro’s row-wise apply vs. rspiro’s vectorised routines) as much as equation arithmetic. At clinically relevant cohort sizes (up to ~10,000 patients), all three packages complete in about one second or less.

### 3.5 Clinical scenario example

To show how the equations and classifiers work together in practice, we built a synthetic patient with moderate COPD and ran a single short script that carries the patient from raw spirometry through to clinical staging. Predicted values and z-scores were first derived with GLI-2012, and the resulting measurements were then handed to the COPD-oriented classifiers. The same record is read as an obstructive pattern of moderate severity, GOLD stage 2, GOLD group E, and a BODE score consistent with that picture, all without leaving Python or stitching separate tools together. The full output is shown in **Table S8**.

### 3.6 Cross-equation comparison

PySpiro leverages its large reference equation database to enable direct comparisons of how different reference equations interpret the same measurement. Given a patient record and a set of equations, the compare_equations() function returns a DataFrame with one row per equation, reporting the percent predicted, z-score, and an applicability flag. Parameters are matched by physiological identity rather than by their internal encoding, so a given parameter is compared consistently even where the equations label it differently. This lets researchers quantify how sensitive a patient’s classification is to the choice of equation, a question with direct clinical relevance, especially when legacy equations are applied to populations they were never derived from.

A worked example makes the effect concrete (**Table S9, Figure 4**). For a hypothetical 40-year-old Caucasian man (175 cm, FEV_1_ 3.2 L), the five equations span a z-score range from −1.18 (BOWERMAN_2022) to −1.89 (HANKINSON_1999), while the pre-LMS polynomial equation KUSTER_2008 yields a percent predicted but no z-score. More tellingly, the interpretation flips: the same 3.2 L is read as below the lower limit of normal under GLI_2012, HANKINSON_1999, and KUSTER_2008, but within the normal range under BOWERMAN_2022 and KUBOTA_2014. The choice of reference equation alone is therefore enough to move this measurement across a clinically meaningful threshold.

**Figure 4.**
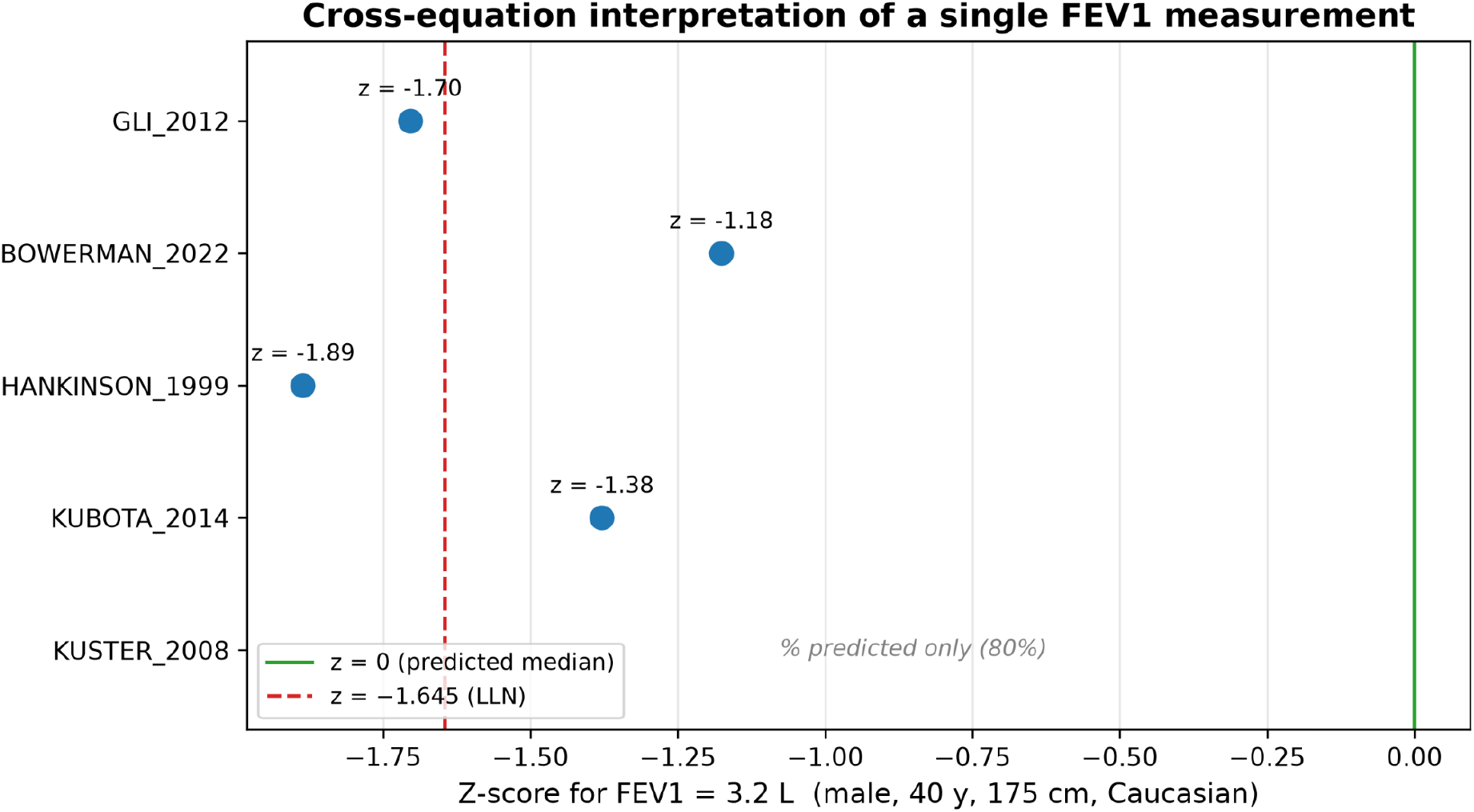
Cross-equation comparison for a single hypothetical input (male, 40 years, 175 cm, Caucasian, measured FEV_1_ 3.2 L). The plot shows the FEV_1_ z-score returned by each reference equation, with the z-score = 0 line (predicted median) and the lower limit of normal (z = −1.645) marked for reference. The pre-LMS polynomial equation KUSTER_2008 returns a percent predicted but no z-score. The same measurement is classified below the lower limit of normal by some equations and within the normal range by others, illustrating the effect of equation choice.

### 3.7 Centile chart visualisation

For the LMS-based equations, PySpiro can also draw the underlying percentile curves across the full validated age range, which makes the differences between equations easy to see at a glance. **Figure 5** plots FEV_1_ centile curves for a 175 cm male under GLI-2012 and the race-neutral BOWERMAN_2022 equation, where the offset between the multi-ethnic and race-neutral references is immediately apparent. The figure is returned as a standard matplotlib object, so it can be restyled or exported for publication directly.

**Figure 5.**
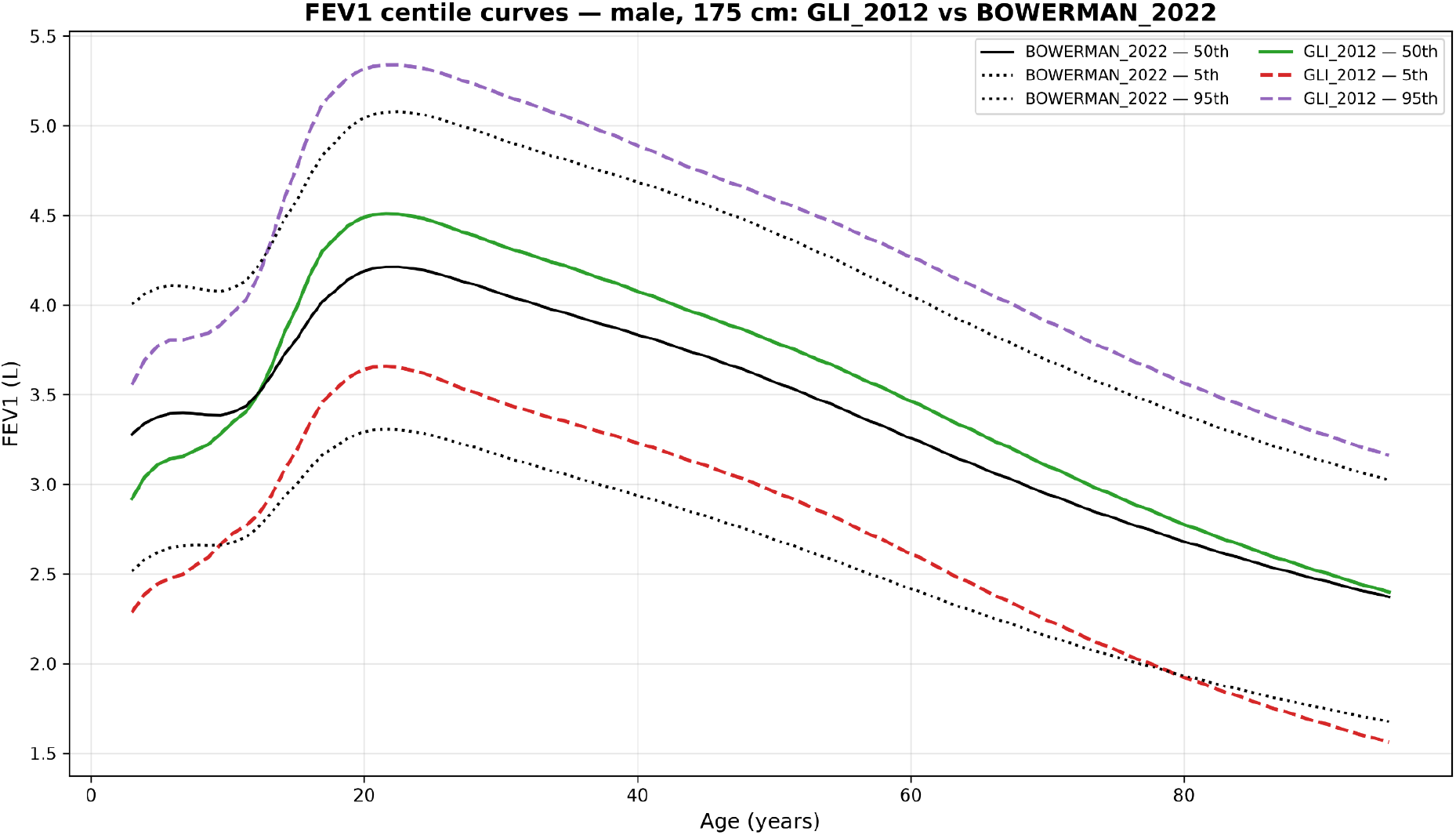
FEV_1_ centile curves across the validated age range for a 175 cm male, computed with the multi-ethnic GLI-2012 and the race-neutral BOWERMAN_2022 equations. Each curve traces a predicted percentile as a function of age; the offset between the two equations shows how the reference set shifts the predicted distribution for the same subject. Curves are produced by plot_centile_curves() function.

## 4. Discussion

PySpiro fills a gap in the respiratory research toolchain by providing, for the first time in Python, a comprehensive, multi-modality, multi-equation library with integrated clinical classifiers.

To our knowledge, there is no other package with that breadth of equation coverage. With 23 spirometry equations spanning 1971-2022, PySpiro covers not only the current gold-standard equations but also legacy equations essential for longitudinal research and equations for populations underserved in existing tools: three Indian, two Korean, one Japanese, and one Chinese equation.

Furthermore, PySpiro provides multi-modality support. No existing Python tool, and no R tool to our knowledge, covers spirometry, MBW, DLCO, static lung volumes, and oscillometry under a single API. This is particularly relevant as modern research protocols increasingly combine modalities. Uniquely, SCAPIS_2023 provides post-bronchodilator reference values for both spirometry and diffusing capacity. This capability is absent from every other equation in PySpiro as well as from rspiro and spiref, and it allows post-bronchodilator measurements to be interpreted against matched reference data rather than pre-bronchodilator norms.

Moreover, since clinical staging requires not only reference values but also classification logic, we integrated multiple classifiers based on clinical guidelines. By co-implementing 10 classifiers in the same package, PySpiro enables seamless downstream analysis without requiring separate tools or manual threshold application.

Beneficially, PySpiro integrates into the Python ecosystem. PySpiro is built on pandas, the standard data manipulation library in Python research pipelines. The compute() method accepts DataFrames directly and returns DataFrames, enabling natural integration with cohort-level analyses, Jupyter notebooks, and machine-learning workflows.

While we already provide a broad range of reference equations, some ethnic groups (Middle Eastern, Sub-Saharan African, South American) are not yet covered. We encourage contributions and requests via the GitHub repository.

## 5. Conclusion

To our knowledge, PySpiro currently provides one of the broadest collections of lung function reference equations available in Python. It provides a validated, extensible platform for researchers working with spirometry, MBW, DLCO, static lung volumes, and oscillometry data, and reduces the barrier to implementing standardised lung function analysis in Python-based research pipelines while providing unrestricted open-source usage and licensing.

## Data Availability

Zenodo DOI: 10.5281/zenodo.15519193
Github: https://github.com/OnlineArts/PySpiro

## Contributions

**Hendrik Pott**: Conceptualization; Methodology; Software; Validation; Formal analysis; Investigation; Resources; Data Curation; Visualization; Writing - Original Draft; Writing - Review & Editing. **Bernd Schmeck**: Funding Acquisition; Project Administration; Supervision; Writing - Original Draft; Writing - Review & Editing. **Dominik Heider**: Funding Acquisition; Project Administration; Validation; Supervision; Writing - Original Draft; Writing - Review & Editing. **Roman Martin**: Conceptualization; Methodology; Software; Validation; Formal analysis; Investigation; Resources; Data Curation; Supervision; Visualization; Writing Original Draft; Writing - Review & Editing.

## Ethics statement

This study used only publicly available, fully de-identified data from the National Health and Nutrition Examination Survey (NHANES) 2011–2012, collected by the U.S. National Center for Health Statistics (NCHS) under its own institutional review board approval and participant consent. No new data were collected and no identifiable participant information was accessed, so no additional ethical approval was required for the present work.

## Highlights

- First comprehensive Python library for lung function reference equations
- Unifies 29 equations across five modalities under a single, consistent API
- Includes ten clinical severity classifiers absent from existing tools
- Numerically cross-validated against rspiro on synthetic and real NHANES data
- Freely available open-source via PyPI with an archived Zenodo DOI

## Funding

This research was funded by the German Federal Ministry of Research, Technology and Space (BMFTR) under the project PerMed-COPD (grant number 01EK2203F).

## Conflicts of interest

The authors declare that they have no competing interests.

## Data availability

Validation used the publicly available, de-identified NHANES 2011–2012 dataset; no new data were collected and no additional ethical approval was required. All reference equation coefficients are encoded from the original publications and stored in the PySpiro repository. The software is available at https://github.com/OnlineArts/PySpiro (doi:10.5281/zenodo.15519193) and can be installed from the PyPI repository.

## Declaration of generative AI use

During the preparation of this work the authors used Claude Opus for grammar and spelling correction, and Claude Code for code polishing and unit-test generation. After using these tools, the authors reviewed and edited the content as needed and took full responsibility for the content of the published article.

## Notes

### Competing Interest Statement

The authors have declared no competing interest.

### Author Declarations

NHANES and simulated data

