## Supplementary Information for "PySpiro: A Python Library for Lung Function Reference Equations and Severity Classification"

### Methods

#### S1 Implementation

All reference equations were implemented directly from their primary publications. Equation coefficients were encoded from the original papers or supplementary materials. For LMS-based (GAMLSS) equations (GLI_2012, BOWERMAN_2022, KUBOTA_2014, JO_2018, JIAN_2017, GLI_2017, GLI_2021, RAMSEY_2024; AGARWAL_2020 uses a pre-computed GAMLSS lookup table), spline coefficients for the Lambda (L), Mu (M), and Sigma (S) parameters are stored as CSV files and loaded at class instantiation. Linear interpolation is performed at non-integer ages and, where applicable, at non-integer heights. For polynomial regression equations (pre-LMS era: MORRIS_1971_1973 through QUANJER_1995), coefficients are encoded directly. Where polynomial-era equations publish a residual spread term (a residual standard deviation, labelled SEE or SE in the source papers), the lower limit of normal is computed as predicted − 1.645 × spread; the z-score uses the same term. HANKINSON_1999 and KUSTER_2008 instead use the lower-limit-of-normal equation given in their original publications (KUSTER_2008 publishes no spread term and therefore returns no z-score). For equations without published limits of normal, `lln()`, `uln()`, and `zscore()` return `pandas.NA`.

The unified scalar API is defined in a base `Reference` class, with `LMSReference` and `SplineReference` subclasses for GAMLSS/LMS and spline-based equations:

- `percent(sex, age, height, ..., parameter, value)` → % of predicted median
- `zscore(sex, age, height, ..., parameter, value)` → z-score
- `lln(sex, age, height, ..., parameter, value)` → 5th-percentile LLN
- `uln(sex, age, height, ..., parameter, value)` → 95th-percentile ULN
- `lms(sex, age, height, ..., parameter, value)` → (L, M, S) triplet (LMS equations only)
- `all(...)` → (% predicted, z-score, LLN, ULN) tuple (LMS/spline equations)
- `compute(df, parameter, ...)` → vectorised apply over a pandas DataFrame

Out-of-range inputs (age or height outside the validated range) return `pandas.NA`. Boundary handling uses a configurable strategy, set via `set_strategy()`: `ignore` (default; return `pandas.NA`) or `closest` (clamp to the nearest range boundary). Out-of-range warning messages can be toggled independently.

Severity classifiers inherit from a `Classifier` base class providing:

- `classify(**kwargs)` → stage label (string) or `pandas.NA`
- `get_order()` → ordered list of stages

Classifiers with a numeric index (e.g. BODE, GAP, ECOPD_ROME_2021) additionally expose a `score(**kwargs)` method returning the raw score.

#### S2 Numerical cross-validation against rspiro

To confirm that PySpiro reproduces an independent reference implementation, we compared it against the rspiro R package for every shared equation. A synthetic cohort of 5000 patients (ages 18–80 y, adult heights, ethnicities 1–4 for GLI / 1–3 for NHANES III) was generated, and z-scores and predicted medians were computed independently by both packages for GLI-2012, GLI-2022, NHANES III, JRS-2014 (FEV1, FVC and, for JRS-2014, VC), and the GLI-2017 diffusing-capacity equation (TLCO, KCO). We additionally compared the Python package spiref against both PySpiro and rspiro for the metrics it provides, predicted values for GLI-2012 and NHANES III, and LLN for GLI-2012; spiref computes no z-scores, so it is restricted to the predicted-value comparison (**Table S3)**. Agreement was assessed by Bland–Altman analysis (**Figure S1**).

Agreement was excellent across all equations. For the LMS/GAMLSS equations the two implementations agreed to floating-point and spline-interpolation precision: JRS-2014 matched to ~$10^{-15}$ (machine epsilon), and the GLI-2012, GLI-2022 and GLI-2017 z-scores agreed with mean bias < $5\times10^{-3}$ and 95% limits of agreement within ±0.02 z-units. The slightly larger residuals for the NHANES III polynomial were confined to the child/adult coefficient transition, where PySpiro and rspiro select the switch age slightly differently; bias remained essentially zero and, away from this boundary, agreement was well within 0.1 z-units. spiref's predicted medians likewise agreed with both PySpiro and rspiro (**Table S3**): for NHANES III, spiref and rspiro matched to machine precision (both evaluating the Hankinson polynomial directly), while the small GLI-2012 residuals reflect spiref's use of floor age-binning of the spline lookup rather than interpolation. The full agreement statistics are reproduced by `python publication/generate_comparison.py` (which invokes rspiro through `rspiro_reference.R`).

### S3 Validation on real NHANES patient data

To confirm the synthetic-cohort agreement on real-world measurements, we repeated the PySpiro vs. rspiro comparison on the NHANES 2011–2012 cohort. Demographic, body-measure and spirometry files were merged on participant identifier; 6309 participants with an acceptable spirogram, a valid standing height, and an ethnicity matching the equation (GLI-2012: Non-Hispanic White or Non-Hispanic Black; NHANES III: additionally Mexican-American) were retained, and GLI-2012 and NHANES III (Hankinson) z-scores and predicted medians were computed independently by both packages.

Agreement on real data matched the synthetic result. For NHANES III, 4196 of 4275 evaluations (98.2%) were bit-identical between the two packages (absolute difference $< 10^{-9}$); the 79 exceptions fell entirely in the child/adult transition band, where PySpiro and rspiro switch coefficient sets at slightly different ages (PySpiro treats females as adult from age 18 and males from age 20, whereas rspiro applies a single age-20 cut-off for both sexes, so only female participants aged 18–19 y fall in the mismatch band; all 79 such participants are female), and away from this band agreement was exact to floating point. Within this band the effect is concentrated at age 19: the child and adult Hankinson coefficient sets for females cross at approximately age 18, so 18-year-old females fall almost exactly on the crossover and agree to within $2\times10^{-4}$ L despite the differing coefficient path, whereas 19-year-old females (the 43 evaluations exceeding $10^{-3}$) differ by up to 0.08 L in predicted FEV1 (about 0.24 z-units). GLI-2012 values agreed to spline-interpolation rounding (largest absolute difference $6\times10^{-4}$). A secondary runtime benchmark on this cohort confirmed that both packages compute the full set of reference values for all 6309 participants in roughly one second. Full statistics are produced by `python publication/generate_nhanes_validation.py`.

### Tables

**Table S1a.** Overview of implemented spirometry reference equations in PySpiro, spiref and rspiro.

| **Equation** | **PySpiro** | **spiref** | **rspiro** |
|---|---|---|---|
| GLI_2012 (Quanjer 2012) multi-ethnic | ✓ | ✓ | ✓ |
| BOWERMAN_2022 (race-neutral GLI global) | ✓ | — | ✓ |
| HANKINSON_1999 (NHANES III) | ✓ | ✓ | ✓ |
| KUBOTA_2014 (Japanese JRS) | ✓ | — | ✓ |
| ECCS_1993 / QUANJER_1993 (ERS/ECCS) | ✓ | — | — |
| CRAPO_1981 (Caucasian US) | ✓ | — | — |
| KNUDSON_1983 (Caucasian US) | ✓ | — | — |
| ROBERTS_1991 (White UK) | ✓ | — | — |
| KUSTER_2008 (Swiss LuftiBus) | ✓ | — | — |
| MORRIS_1971_1973 (Caucasian US) | ✓ | — | — |
| CHERNIACK_1972 (Caucasian US) | ✓ | — | — |
| POLGAR_1971 (children) | ✓ | — | — |
| WARWICK_1980 (US children) | ✓ | — | — |
| QUANJER_1995 (European children) | ✓ | — | — |
| WANG_1993 (White & African-American children, US children) | ✓ | — | — |
| ZAPLETAL_1987 (European children) | ✓ | — | — |
| HSU_1979 (White, African-American, Mexican-American children) | ✓ | — | — |
| AGARWAL_2020 (Western Indian) | ✓ | — | — |
| CHHABRA_2014 (Northern Indian) | ✓ | — | — |
| DESAI_2016 (Western Indian, Mumbai) | ✓ | — | — |
| JIAN_2017 (Han Chinese) | ✓ | — | — |
| JO_2018 (Korean KNHANES IV–V) | ✓ | — | — |
| CHOI_2005 (Korean KNHANES 2001–02) | ✓ | — | — |
| **Total spirometry equations** | **23** | **2** | **4** |

**Table S1b.** Overview of different modalities and equations implemented in PySpiro, spiref, and rspiro.

| **Modality / Equation** | **PySpiro** | **spiref** | **rspiro** |
|---|---|---|---|
| Multiple breath washout: RAMSEY_2024 (GLI, LCI + FRC) | ✓ | — | — |
| Transfer factor: GLI_2017 (DLCO) | ✓ | — | ✓ |
| Pre-/post-bronchodilator reference values (DLCO, $FEV_1$) | ✓ | — | — |
| Static lung volumes: GLI_2021 (TLC, RV, FRC, VC) | ✓ | — | — |
| Oscillometry: SCHULZ_2013 (IOS, adults) | ✓ | — | — |
| Oscillometry: CALOGERO_2013 (IOS, children) | ✓ | — | — |
| **Modality besides spirometry** | **6** | **0** | **1** |

**Table S1c.** Overview of features available in PySpiro, spiref and rspiro. (*) vectorised.

| **Features** | **PySpiro** | **spiref** | **rspiro** |
|---|---|---|---|
| Predicted median | ✓ | ✓ | ✓ |
| % of predicted | ✓ | — | ✓ |
| z-scores | ✓ | — | ✓ |
| LLN (5th percentile) | ✓ | ✓ | ✓ |
| ULN (95th percentile) | ✓ | — | — |
| Raw LMS triplet | ✓ | — | ✓ |
| All metrics in one call | ✓ | — | — |
| DataFrame batch processing | ✓ | — | ✓ * |
| Configurable out-of-range strategy | ✓ | — | — |
| Centile curve visualisation | ✓ | — | — |
| Cross-equation comparison table | ✓ | — | — |
| **Total features** | **11** | **2** | **6** |

**Table S1d.** Programming language, licensing, latest release and package manager of PySpiro, spiref and rspiro.

| **Feature** | **PySpiro** | **spiref** | **rspiro** |
|---|---|---|---|
| Language | Python | Python | R |
| Licence | MIT | MIT | GPL (≥2) |
| Latest release | 2025 (doi:10.5281/zenodo.15519193) | 2020 (v1.0.4) | 2024 (v0.5) |
| Package manager | PyPI | PyPI | CRAN |

**Table S2.** Validation of PySpiro against published reference values.

| Equation | Synthetic input | Parameter | Measured | Predicted (M) | % predicted | z-score | LLN | Published value |
|---|---|---|---|---|---|---|---|---|
| GLI_2012 | M 40y 175cm Caucasian | FEV1 | 3.0 L | 4.078 L | 73.6 | -2.08 | 3.231 L | 73.6% (Quanjer 2012) |
| GLI_2012 | M 40y 175cm Caucasian | FEV1 LLN | 3.231 L | — | — | -1.645 | — | z = −1.645 (definition) |
| BOWERMAN_2022 | M 40y 175cm | FVC | 4.5 L | 4.699 L | 95.8 | -0.30 | 3.618 L | 95.77% (Bowerman 2023) |
| GLI_2017 | M 40y 175cm | KCO (SI) | 1.5 mmol/min/kPa/L | 1.585 | 94.6 | -0.41 | — | 94.62% (Stanojevic 2017) |
| RAMSEY_2024 | any, age 5 | LCI ULN | — | — | — | — | — | ULN ≈ 7.52 (Ramsey 2024 Table 3); computed: 7.52 |
| RAMSEY_2024 | any, age 20 | LCI ULN | — | — | — | — | — | ULN ≈ 6.85 (Ramsey 2024 Table 3); computed: 6.85 |
| HANKINSON_1999 | M 40y 175cm Caucasian | FEV1 | 3.5 L | 4.075 L | 85.9 | -1.24 | 3.312 L | 4.075 L (Hankinson 1999 Table 4 coefficients) |

**Table S3a. Agreement of z-scores between PySpiro and rspiro across the shared reference equations (5000 synthetic patients, ages 18–80 y).** For each equation and parameter the per-patient difference (PySpiro − rspiro) is summarised by its mean (bias), standard deviation (SD), 95% limits of agreement (bias ± 1.96 SD), and maximum absolute difference. NHANES III is restricted to the three ethnicities it defines (n = 3627); the LMS equations cover all 5000 subjects.

| **Equation / parameter** | **n** | **Bias** | **SD** | **95% limits of agreement** | **Max \|Δ\|** |
|---|---|---|---|---|---|
| GLI-2012 FEV1 | 5000 | +4.88e-05 | 4.84e-03 | -9.45e-03 to +9.54e-03 | 1.56e-02 |
| GLI-2012 FVC | 5000 | +3.25e-05 | 3.70e-03 | -7.22e-03 to +7.29e-03 | 2.83e-02 |
| GLI-2022 FEV1 | 5000 | +4.22e-03 | 7.55e-03 | -1.06e-02 to +1.90e-02 | 3.79e-02 |
| GLI-2022 FVC | 5000 | +2.65e-03 | 5.74e-03 | -8.60e-03 to +1.39e-02 | 3.35e-02 |
| NHANES III FEV1 | 3627 | +3.35e-03 | 3.62e-02 | -6.76e-02 to +7.43e-02 | 4.42e-01 |
| NHANES III FVC | 3627 | +2.62e-03 | 3.19e-02 | -5.99e-02 to +6.52e-02 | 4.17e-01 |
| JRS-2014 FEV1 | 5000 | +5.00e-17 | 2.39e-15 | -4.63e-15 to +4.73e-15 | 8.88e-15 |
| JRS-2014 FVC | 5000 | -6.08e-18 | 2.31e-15 | -4.52e-15 to +4.51e-15 | 6.66e-15 |
| JRS-2014 VC | 5000 | -4.50e-17 | 2.34e-15 | -4.62e-15 to +4.53e-15 | 5.33e-15 |
| GLI-2017 TLCO | 5000 | +1.88e-03 | 4.83e-03 | -7.59e-03 to +1.14e-02 | 3.05e-02 |
| GLI-2017 KCO | 5000 | +6.76e-04 | 2.98e-03 | -5.16e-03 to +6.51e-03 | 1.65e-02 |

**Table S3b. Agreement of predicted medians between PySpiro and rspiro.** Same cohort and layout as **Table S3a.** Predicted medians are independent of the measured value and so isolate the core equation arithmetic from the z-score transform.

| **Equation / parameter** | **n** | **Bias** | **SD** | **95% limits of agreement** | **Max \|Δ\|** |
|---|---|---|---|---|---|
| GLI-2012 FEV1 | 5000 | -2.03e-05 | 1.88e-03 | -3.70e-03 to +3.66e-03 | 8.04e-03 |
| GLI-2012 FVC | 5000 | -2.33e-05 | 1.88e-03 | -3.72e-03 to +3.67e-03 | 1.09e-02 |
| GLI-2022 FEV1 | 5000 | -1.82e-03 | 3.32e-03 | -8.33e-03 to +4.68e-03 | 1.94e-02 |
| GLI-2022 FVC | 5000 | -1.54e-03 | 3.40e-03 | -8.19e-03 to +5.12e-03 | 1.97e-02 |
| NHANES III FEV1 | 3627 | -1.14e-03 | 1.39e-02 | -2.85e-02 to +2.62e-02 | 1.54e-01 |
| NHANES III FVC | 3627 | -1.03e-03 | 1.48e-02 | -3.00e-02 to +2.79e-02 | 1.79e-01 |
| JRS-2014 FEV1 | 5000 | -1.69e-16 | 2.87e-15 | -5.80e-15 to +5.46e-15 | 5.33e-15 |
| JRS-2014 FVC | 5000 | -1.02e-16 | 2.93e-15 | -5.84e-15 to +5.64e-15 | 5.33e-15 |
| JRS-2014 VC | 5000 | +4.81e-17 | 2.91e-15 | -5.66e-15 to +5.75e-15 | 5.33e-15 |
| GLI-2017 TLCO | 5000 | -2.66e-03 | 6.65e-03 | -1.57e-02 to +1.04e-02 | 4.49e-02 |
| GLI-2017 KCO | 5000 | -1.77e-04 | 6.15e-04 | -1.38e-03 to +1.03e-03 | 2.63e-03 |

**Table S3c. Agreement of spiref predicted values against PySpiro and rspiro (same cohort).** spiref (PyPI 1.0.4) computes predicted values for GLI-2012 and NHANES III and LLN for GLI-2012 only, no z-scores, % predicted, ULN or DataFrame interface, so it is restricted to the predicted-value comparison and is absent from **Tables S3a and S3b**. Bias and maximum absolute difference are shown against each of the other two packages.

| **Metric / parameter** | **n** | **spiref−PySpiro bias** | **max \|Δ\|** | **spiref−rspiro bias** | **max \|Δ\|** |
|---|---|---|---|---|---|
| GLI-2012 FEV1 predicted | 5000 | +2.52e-03 | 1.66e-02 | +2.50e-03 | 1.27e-02 |
| GLI-2012 FVC predicted | 5000 | +2.59e-03 | 2.28e-02 | +2.57e-03 | 1.68e-02 |
| NHANES III FEV1 predicted | 3627 | +1.14e-03 | 1.54e-01 | -1.26e-16 | 1.78e-15 |
| NHANES III FVC predicted | 3627 | +1.03e-03 | 1.79e-01 | -1.13e-16 | 2.66e-15 |
| GLI-2012 FEV1 LLN | 5000 | +2.27e-03 | 1.72e-02 | — | — |
| GLI-2012 FVC LLN | 5000 | +2.37e-03 | 2.05e-02 | — | — |

**Table S4. Numerical agreement between PySpiro and rspiro on real NHANES 2011–2012 participants.** For each equation, parameter, and metric the table reports the maximum and mean absolute difference (PySpiro minus rspiro), the number of participants for whom the two packages were bit-identical (absolute difference $< 10^{-9}$), and the number agreeing to within $10^{-3}$. GLI-2012 values never coincide bit-for-bit, because rspiro evaluates its spline on coefficients rounded to four decimal places whereas PySpiro retains full precision, but all participants agree to within $10^{-3}$. NHANES III values are bit-identical for 4196 of 4275 participants; the 79 exceptions are all female participants aged 18–19 y, for whom PySpiro and rspiro apply the child/adult coefficient cut-off at different ages. Of these, only the 43 aged 19 y exceed the $10^{-3}$ threshold; the 36 aged 18 y agree to within $2\times10^{-4}$ because age 18 lies almost exactly on the child/adult crossover of the female Hankinson curves.

| Comparison | n | max \|Δ\| | mean \|Δ\| | n bit-identical ($<10^{-9}$) | n within $10^{-3}$ |
|---|---:|---:|---:|---:|---:|
| GLI-2012 FEV1 z-score | 3708 | 6.12e-04 | 1.92e-04 | 0/3708 | 3708/3708 |
| GLI-2012 FEV1 predicted | 3708 | 2.44e-04 | 7.67e-05 | 0/3708 | 3708/3708 |
| GLI-2012 FVC z-score | 3708 | 5.42e-04 | 1.77e-04 | 0/3708 | 3708/3708 |
| GLI-2012 FVC predicted | 3708 | 3.07e-04 | 8.71e-05 | 0/3708 | 3708/3708 |
| NHANES III FEV1 z-score | 4275 | 2.42e-01 | 2.06e-03 | 4196/4275 | 4232/4275 |
| NHANES III FEV1 predicted | 4275 | 8.07e-02 | 7.63e-04 | 4196/4275 | 4232/4275 |
| NHANES III FVC z-score | 4275 | 2.45e-01 | 1.88e-03 | 4196/4275 | 4232/4275 |
| NHANES III FVC predicted | 4275 | 9.09e-02 | 8.22e-04 | 4196/4275 | 4232/4275 |

**Table S5a. PySpiro `compute()` z-score time (milliseconds) by cohort size**, median of three runs (Python 3.12, single core; NumPy 2.3, pandas 2.3). Benchmarked on an AMD Ryzen 5 Pro 7535U notebook CPU. Each cohort is a prefix of the largest, so all sizes and both packages (**Table S5b**) are timed on identical patients.

| Equation | 100 | 300 | 1,000 | 3,000 | 10,000 | 30,000 | 100,000 |
|---|---|---|---|---|---|---|---|
| GLI-2012 (LMS spline) | 10.4 | 28.6 | 93.3 | 281.6 | 941.5 | 2855.9 | 9888.0 |
| GLI-2022 (LMS spline) | 7.4 | 21.0 | 69.0 | 204.4 | 682.0 | 2025.8 | 6827.3 |
| NHANES III (polynomial) | 7.4 | 21.9 | 68.4 | 206.1 | 676.8 | 2048.9 | 6765.6 |
| JRS-2014 (LMS) | 9.8 | 28.9 | 96.0 | 291.4 | 945.0 | 2844.8 | 9566.7 |
| GLI-2017 (LMS spline) | 7.6 | 21.6 | 70.0 | 213.3 | 700.6 | 2056.4 | 6834.4 |

**Table S5b. rspiro z-score time (milliseconds) by cohort size** on the same patients (median of three runs).

| Equation | 100 | 300 | 1,000 | 3,000 | 10,000 | 30,000 | 100,000 |
|---|---|---|---|---|---|---|---|
| GLI-2012 | 9.76 | 13.61 | 28.08 | 70.53 | 240.48 | 773.71 | 2935.83 |
| GLI-2022 | 7.48 | 10.44 | 21.59 | 55.17 | 176.12 | 609.10 | 2552.67 |
| NHANES III | 8.19 | 22.72 | 81.63 | 253.84 | 768.85 | 2179.90 | 7351.67 |
| JRS-2014 | 5.15 | 7.85 | 18.58 | 50.18 | 165.77 | 545.95 | 2339.67 |
| GLI-2017 | 6.80 | 9.34 | 19.91 | 51.30 | 166.82 | 551.95 | 2189.83 |

**Table S6. Z-score throughput at n = 100,000 (patients per second)** for PySpiro and rspiro, with the rspiro-to-PySpiro speed ratio.

| Equation | PySpiro | rspiro | rspiro / PySpiro |
|---|---|---|---|
| GLI-2012 | 10,113 | 34,062 | 3.4× |
| GLI-2022 | 14,647 | 39,175 | 2.7× |
| NHANES III | 14,781 | 13,602 | 0.9× |
| JRS-2014 | 10,453 | 42,741 | 4.1× |
| GLI-2017 | 14,632 | 45,666 | 3.1× |

**Table S7. Predicted-value (FEV1) throughput at n=100,000 (patients per second)** for the predicted-value task, the only operation spiref supports, across all three packages. PySpiro evaluates the predicted median row-wise (pandas `apply`), spiref via a Python list comprehension, rspiro via its vectorised `pred_*` functions; the figures therefore index call style as much as equation arithmetic and should be read as order-of-magnitude throughput.

| Equation | PySpiro | rspiro | spiref |
|---|---|---|---|
| GLI-2012 FEV1 | 9,873 | 50,268 | 319,359 |
| NHANES III FEV1 | 14,168 | 15,635 | 1,589,164 |

**Table S8a.** Spirometry GLI_2012; hypothetical (fictitious) clinical scenario: 63-year-old male with COPD. Height 172 cm; post-BD $FEV_1$ = 1.8 L, FVC = 3.6 L; 6MWT = 280 m; mMRC = 2; BMI = 26.3; 2 moderate exacerbations in the past 12 months; CAT = 18.

| Parameter | Measured | Predicted | % predicted | z-score |
|---|---|---|---|---|
| FEV1 | 1.8 L | 3.23 L | 55.7 | -2.77 |
| FVC | 3.6 L | 4.18 L | 86.2 | -0.93 |
| FEV1/FVC | 0.50 | 0.77 | 64.6 | -3.28 |

**Table S8b.** Classifier output based on the same hypothetical input data from **Table S8a**.

| Module | Classifier | Result |
|---|---|---|
| ATS_ERS_2022 | ATS/ERS 2022 pattern | Obstructive |
| LF_SEVERITY_2022 | LF severity (FEV1) | Moderate |
| GOLD | GOLD spirometric stage | 2 |
| GOLD_ABE | GOLD ABE group | E |
| BODE | BODE index | score 3, quartile 2 |
| ECOPD_ROME_2021 | ECOPD severity (Rome 2021) | Moderate (abnormal 5/5, assessed 5) |

**Table S9.** Cross-equation comparison: FEV1 for a hypothetical input (40-year-old male Caucasian: Height 175 cm, measured $FEV_1$ = 3.2 L).

| Equation | Predicted (L) | % predicted | z-score | LLN (L) | Below LLN |
|---|---|---|---|---|---|
| GLI_2012 | 4.08 | 78.5 | -1.70 | 3.23 | Yes |
| BOWERMAN_2022 | 3.84 | 83.4 | -1.18 | 2.94 | No |
| HANKINSON_1999 | 4.07 | 78.5 | -1.89 | 3.31 | Yes |

| Equation | Predicted (L) | % predicted | z-score | LLN (L) | Below LLN |
|---|---|---|---|---|---|
| KUBOTA_2014 | 3.76 | 85.0 | -1.38 | 3.09 | No |
| KUSTER_2008 | 4.01 | 79.8 | N/A | 3.20 | Yes |

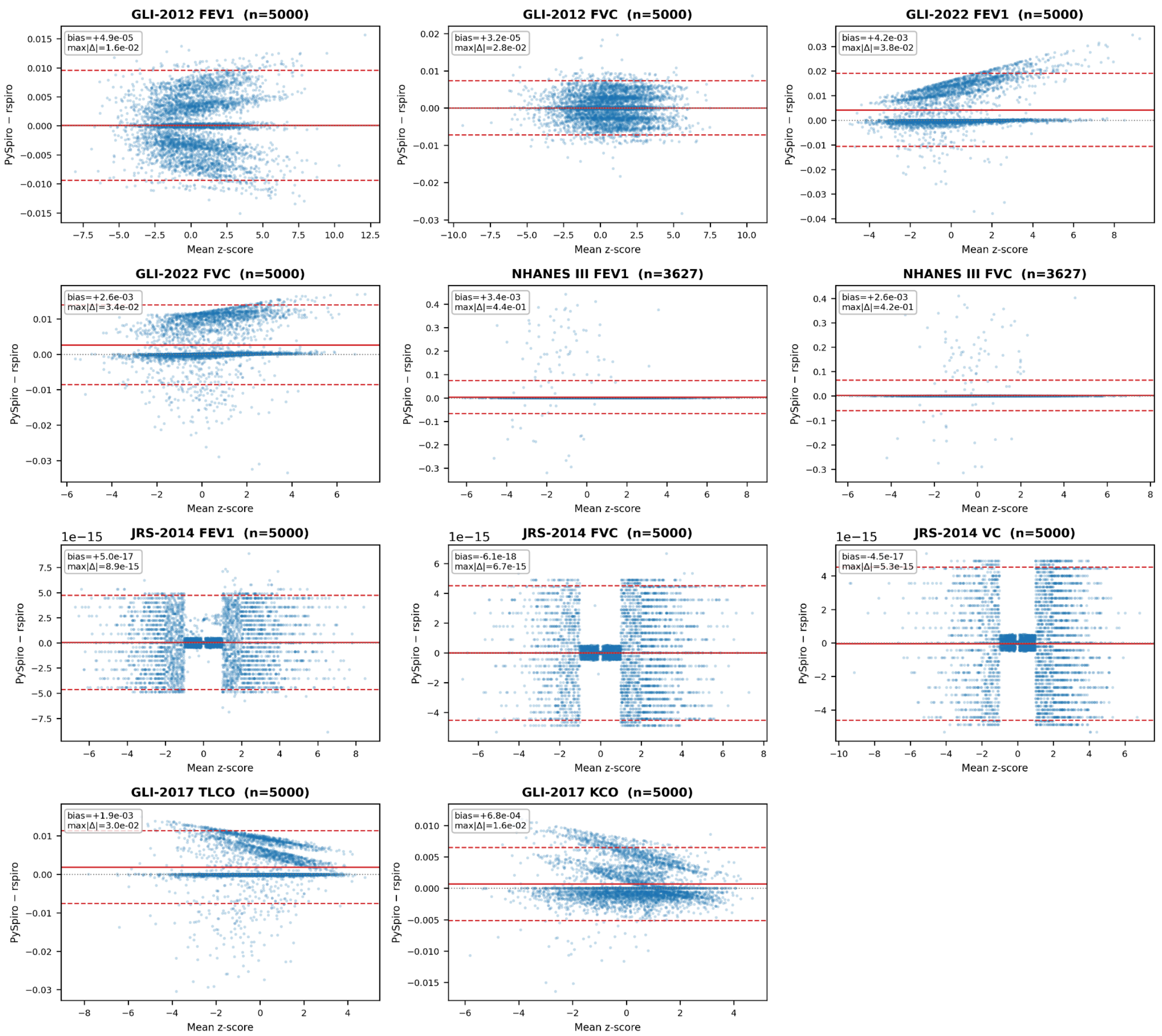


**Figure S1.** Bland–Altman agreement between PySpiro and rspiro z-scores on a synthetic cohort of 5000 simulated records (ages 18 to 80 y). Each of the eleven panels corresponds to one equation and parameter and plots the per-patient z-score difference (PySpiro minus rspiro) on the y-axis against the mean of the two estimates on the x-axis; points are semi-transparent to convey density. In each panel the solid line marks the mean difference (bias), the dashed lines the 95% limits of agreement, and the dotted line zero, with the bias and maximum absolute difference annotated. Note the panel-specific y-axis scales: differences are at the $10^{-2}$ level for the GLI-family LMS equations (spline-interpolation rounding) and at machine-epsilon ($\sim 10^{-15}$) for JRS-2014, and are largest for the NHANES III polynomial, where the few outlying points fall at the child/adult coefficient transition.
